# SOFA-2 reclassifies multiorgan dysfunction syndrome in major trauma patients

**DOI:** 10.64898/2026.08.05.26359771

**Authors:** Anand Krishna, Andrea Rossetto, Karim Brohi, Paul Vulliamy, Elaine Cole

## Abstract

**Objective:** We aimed to evaluate the performance of the recently updated Sequential Organ Failure Assessment Score-2 (SOFA-2) on organ dysfunction classification and prognostication compared to SOFA-1 in critically injured trauma patients.

**Methods:** Adult trauma patients admitted to critical care at four urban Major Trauma Centres between 2011 and 2024 were included. Daily organ dysfunction scoring was performed using SOFA-1 and SOFA-2 until death or discharge. The primary outcome was MODS, defined as SOFA score ≥6.

**Results:** In 2162 severely injured patients (median Injury Severity Score 25 [IQR, 17-34]), SOFA-2 reduced the proportion of patients classified as having MODS compared with SOFA-1 (61.6% vs 68.5%, p<0.001). SOFA-2 scores on the first day after admission were lower than SOFA-1 (median 6 [IQR, 3-8] vs 7 [IQR, 4-10], p<0.001), driven predominantly by lower respiratory and cardiovascular scoring. Critical care mortality in trauma patients was increased in respiratory, cardiovascular and renal components of SOFA-2 at the higher ends of the scores, consistent with the aims of the SOFA-2 reclassification. A group of 159 severely injured patients (7.3%) classified as MODS by SOFA-1 were reclassified to no-MODS by SOFA-2. Despite this reclassification, these patients had substantially higher ICU mortality (7.5% vs 0.7%, p<0.01), greater ventilator and vasopressor requirements, and longer hospital stays than patients classified as no-MODS by both systems.

**Conclusions:** SOFA-2 reduces MODS prevalence in severely injured patients and changes organ dysfunction classification, with lower rates of severe respiratory and cardiovascular dysfunction. This represents an important update in trauma MODS measurement and has implications for future trauma trial design. However SOFA-2 reclassification generates a small cohort a small but clinically significant group with ‘occult MODS’ that warrants further evaluation in severely injured trauma patients.

## Introduction

Multiple organ dysfunction syndrome (MODS) is a primary cause of mortality and morbidity in severely injured trauma patients who survive beyond the initial traumatic insult(1), (2), as well as being resource-intensive for trauma systems (3). The prevalence of MODS in trauma patients varies between 10% to 65%, with critical care mortality up to 40% (4). Whilst MODS is defined using differing scoring systems, the Sequential Organ Failure Assessment score (SOFA), originally developed to measure acute morbidity in patients with critical illness (5,6), is widely used in observational and interventional studies in trauma cohorts (7–11).

Since its original publication in 1996, developments in critical care mean that SOFA may not capture contemporary medications or devices used in patient management (12,13). As a result, the SOFA-2 score was recently developed and validated in 3.3 million adult patients across ten international multicentre cohorts (5). SOFA-2 incorporates contemporary organ support to better define organ dysfunction, and revised thresholds to improve predictive validity for critical care mortality. However the new score was validated predominantly in medical and elective surgical cohorts, and the extent of trauma patient inclusion is uncertain. It is unclear how SOFA-2 reclassification will influence the prevalence, organ contribution and outcomes of trauma-related organ dysfunction (2,14).

The objective of this study was to evaluate the impact of the revised SOFA-2 scoring system on the classification and prognostic assessment of organ dysfunction in trauma patients. Our aims were (1) to compare organ-specific and total SOFA scores between SOFA-1 and SOFA-2 scoring systems; (2) to describe changes in the characteristics of patients scored as having MODS using the update system; (3) to determine the alignment between SOFA-2 scores and outcomes including ICU mortality, organ support and length of stay.

## Materials and methods

This study is reported according to the Strengthening the Reporting of Observational Studies in Epidemiology (STROBE) Statement (**Supplemental Table**).

Patients were included if they had been enrolled in the Activation of Coagulation and Inflammation in Trauma II observational study (ACIT-II, NHS Research Ethics Committee: 07/Q0603/29) (15) between 2011 and 2024 or in the Multiple Organ Dysfunction in Elderly Trauma study (MODET, NHS Research Ethics Committee: 16/LO/1720) (16) between 2017 and 2018. For inclusion, trauma patients had to be aged 16 years or older and admitted to critical care at any point in their hospital stay at one of the four urban Major Trauma Centres (MTCs, Level 1 equivalent hospitals) involved in these studies. Critical care admission was defined as either high dependency unit (HDU, level 2) or intensive care unit (ICU, level 3) care (17). We excluded those who died in the emergency department or operating theatre prior to critical care admission. Detailed study procedures relating to the ACIT-II and MODET studies have been reported previously (15,16).

### Data Collection and Outcomes

Within both studies SOFA scores were collected daily from admission to critical care and patients were followed up until discharge or death to a maximum of 28 days. SOFA allocates scores for organ dysfunction in six individual systems: brain, respiratory, cardiovascular, kidney, liver and haemostasis. Each individual organ system is graded from 0–4 points reflecting the severity of organ dysfunction and failure; these are added together to give a total score ranging from 0-24. For this analysis a score of 0 represented normal function, whilst 1 was mild dysfunction, 2 denoted moderate dysfunction, 3 was severe dysfunction and 4 indicated very severe dysfunction or failure (18). Organ scores were calculated using the original SOFA-1 (19) and SOFA-2 (5). Missing component scores were resolved by allocating the same score as the previous day, according to the methodology used in the updated SOFA-2 scoring system and previous studies of MODS in trauma (14).

Data were collected on demographic variables, injury severity, admission physiology and resuscitation. The primary outcome was MODS, defined as SOFA score ≥ 6 at any point during critical care stay (8). Secondary outcomes were vasopressor days, ventilator days, 28-day mortality, days in critical care and hospital length of stay. Vasopressor days, ventilator days and length of stay variables were calculated for survivors only.

### Data Analysis

Data analysis was conducted using statistical software R (v4.4.1, Vienna, Austria) and RStudio (v2023.9.1.494, Boston, USA) (20,21). Continuous variables were treated as non-parametric, reported as median with interquartile range (IQR) and analysed using the Kruskal-Wallis test with Dunn’s post-hoc test or the Mann-Whitney U-test. with Bonferroni correction when required. Categorical variables were compared using Chi-squared tests and reported as frequency and percentage. Paired tests, such the Wilcoxon signed-rank test and the McNemar test, were used to compare the SOFA score on the first day after admission to hospital, the frequency of MODS according to SOFA-1 or SOFA-2 scoring and the scoring transitions in individual SOFA components. Bonferroni correction was used when required. Previous studies of MODS in trauma populations revealed that organ scores peak at 24 hours in the majority of patients (2,16), and therefore we examined differences between scoring systems on the first day after admission. For reclassification analysis we grouped patients according to MODS status at any time during in-hospital stay: SOFA-1 No MODS, SOFA-2 No MODS, SOFA-1 MODS and SOFA-2 MODS. Survival times between groups were compared using log-rank test and presented as Kaplan–Meier curves.

## Results

There were 2162 patients from both studies with SOFA scores available for analysis (**Supplemental Fig. 1**). The cohort was severely injured, with a median ISS of 25 (IQR 17-34), and the majority (84%) had sustained blunt trauma (**Table 1**). Overall, critical care mortality was 14.5% and in survivors, median critical care stay was 9 days (IQR 4-18). Applying the updated SOFA-2 scoring system resulted in a significant reduction in the proportion of patients classified as developing MODS compared to SOFA-1 (61% vs 69%, p<0.001) (**Table 1**). Patients classified as being in MODS by SOFA-2 were more shocked than those without MODS (Base Deficit: 4.5mmol/L [IQR 1.7-8.3] vs 2.6mmol/L [IQR 0.4-5.1], p<0.001) and had a higher incidence of TBI (56.6% vs 29.4%, p<0.001). This group also had a significantly higher critical care mortality rate (22.3% vs 2.0%, p<0.001) and length of critical care stay (12 days [IQR 6-22] vs 4 days [IQR 3-7], p<0.001) compared to those who did not develop MODS.

**Table 1.** Clinical characteristics according to SOFA-1 and SOFA-2 MODS classification.

|  |  | MODS |  | No MODS |  |
| --- | --- | --- | --- | --- | --- |
|  |  | SOFA-1 | SOFA-2 | SOFA-1 | SOFA-2 |
|  |  | 1480 (68.5) | 1331 (61.6) | 682 (31.5) | 831 (38.4) |
|  | **Demographics** |  |  |  |  |
|  | Age, years | 48 (30-65) | 48 (30-65) | 41 (25-61) | 41 (25-63) |
|  | Sex, male | 1146 (77.5%) | 1037 (78.0%) | 513 (75.2%) | 622 (74.8%) |
|  | **Clinical characteristics** |  |  |  |  |
|  | SBP, mmHg | 124 (98-147) | 124 (97-147) | 125 (102-143) | 125 (103-144) |
|  | Glasgow coma score | 10 (4-14) | 9 (4-14) | 15 (13-15) | 15 (12-15) |
|  | Base deficit, mEq/L | 4.3 (1.4-8.1) | 4.5 (1.7-8.3) | 2.6 (0.4-5.0) | 2.6 (0.4-5.1) |
|  | **Injury characteristics** |  |  |  |  |
|  | MOI, blunt | 1306 (88.2%) | 1192 (89.6%) | 515 (75.5%) | 629 (75.7%) |
|  | Traumatic brain injury | 816 (55.1%) | 753 (56.6%) | 181 (26.5%) | 244 (29.4%) |
|  | Injury severity score | 26 (18-36) | 26 (18-38) | 25 (16-29) | 25 (16-29) |
|  | Head Neck AIS | 3 (0-5) | 3 (0-5) | 1 (0-4) | 2 (0-4) |
|  | Face AIS | 0 (0-1) | 0 (0-1) | 0 (0-0) | 0 (0-0) |
|  | Thorax AIS | 3 (0-4) | 3 (0-4) | 2 (0-3) | 2 (0-3) |
|  | Abdomen-Pelvis AIS | 0 (0-2) | 0 (0-2) | 0 (0-2) | 0 (0-2) |
|  | Extremity AIS | 0 (0-2) | 0 (0-2) | 0 (0-2) | 0 (0-2) |
|  | External AIS | 0 (0-0) | 0 (0-0) | 0 (0-0) | 0 (0-0) |
|  | **Transfusion requirements** |  |  |  |  |
|  | MHP Activation | 586 (40.0%) | 541 (41.0%) | 179 (26.4%) | 224 (27.2%) |
|  | Red blood cells in 24hr | 1 (0-5) | 1 (0-5) | 0 (0-2) | 0 (0-3) |
|  | **Outcomes** |  |  |  |  |
|  | Days in MODS | 5 (2-10) | 4 (2-7) | - | - |
|  | Critical care mortality | 308 (20.8%) | 297 (22.3%) | 6 (0.9%) | 17 (2.0%) |
|  | Critical care length of stay | 12 (6-21) | 12 (6-22) | 4.0 (2.0-6.0) | 4.0 (3.0-7.0) |
|  | 28/7 Mortality | 352 (23.8%) | 340 (25.5%) | 17 (2.5%) | 29 (3.5%) |
|  | Length of stay | 35 (22-52) | 37 (25-55) | 14 (8-24) | 15 (9-26) |

(interquartile range) or frequency (percentage). Days in MODS was calculated according to SOFA-1 and SOFA-2 respectively. MODS, multiorgan dysfunction syndrome; SOFA, sequential organ function assessment; SBP, systolic blood pressure; MOI, mechanism of injury; AIS, abbreviated injury score; MHP, major haemorrhage protocol.

### Comparing SOFA-1 and SOFA-2 organ dysfunction scoring

Median total SOFA-2 scores on the first day of admission were lower than SOFA-1 (SOFA-2: 6 [3-8], SOFA-1: 7 [4-10], p<0.001 **Fig. 1A**). Most of these changes were captured by differences in respiratory and cardiovascular components of the SOFA scores (**Fig. 1B**). For the respiratory system, the proportion of patients classified as having severe dysfunction or failure (score 3-4) fell substantially, from 26.0% in SOFA-1 to 11.8% in SOFA-2 (p<0.001). Similarly, the proportion of patients classified as having severe cardiovascular dysfunction or failure decreased from 50.9% in SOFA-1 to 28.1% in SOFA-2 (p<0.001). There was a corresponding increase in the proportion of patients scored as moderate cardiovascular dysfunction, from 2.4% to 25.2% (p<0.001). For the haemostasis component, SOFA-2 classified 10.3% of patients as severe dysfunction or failure, from only 1.7% with SOFA-1 scoring (p<0.001). There were no major changes in the distribution of scores for the brain, liver or kidney components. Collectively, this shows that SOFA-2 scoring reduces total organ failure scores in trauma patients compared to SOFA-1, primarily driven by altered scores in the respiratory and cardiovascular domains.

**Figure 1.**
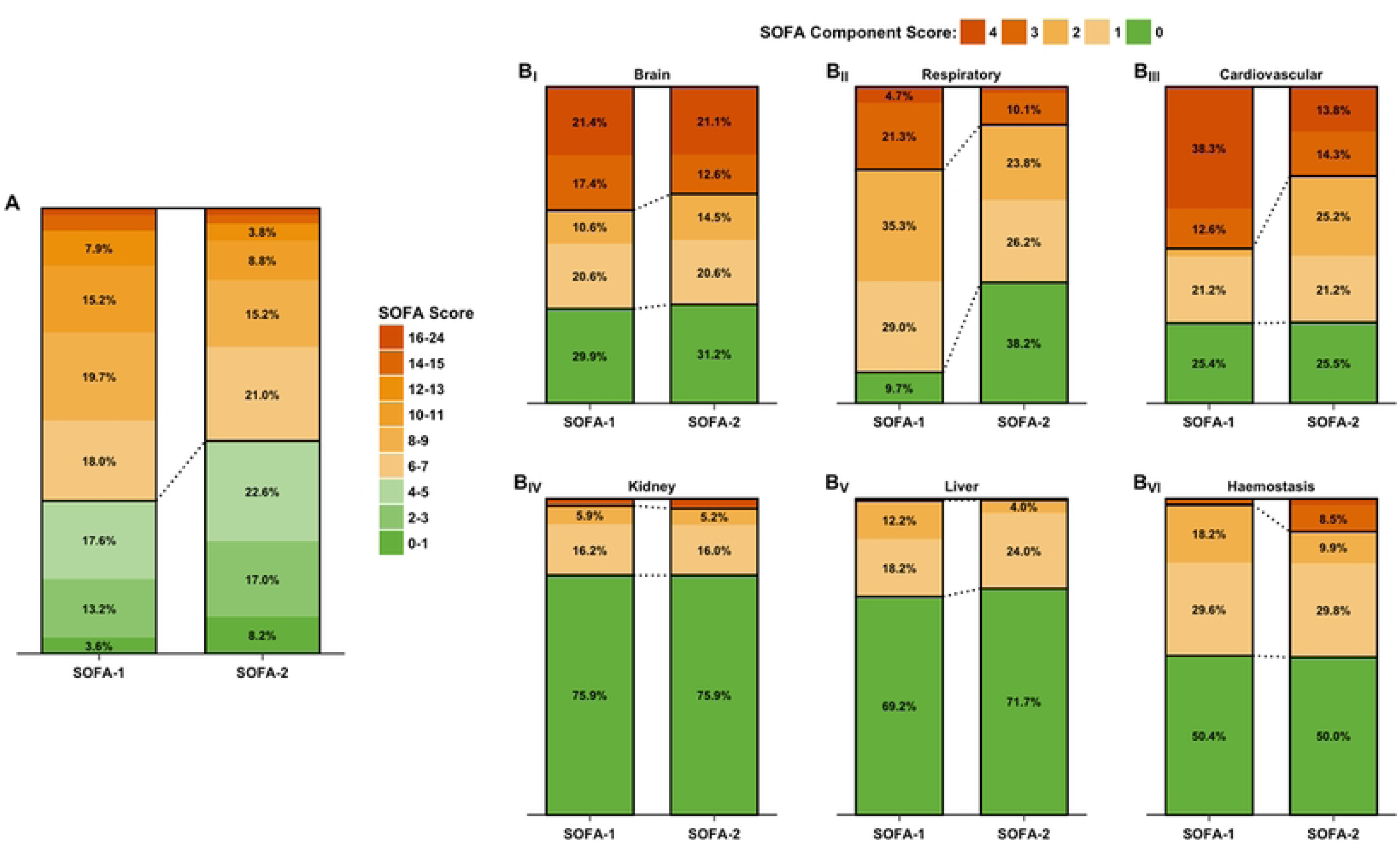
Changes in distribution of total SOFA scores at 24h of critical care admission according to SOFA-1 vs SOFA-2. A: Total SOFA score at 24 hours, B: SOFA Scores in individual organs. Green scale: No-MODS; orange scale: MODS. Proportions not reported in A: SOFA-1 group 14-15 = 3.5%, SOFA-1 group 16-24 = 1.4%, SOFA-2 group 14-15 = 1.9% and SOFA-2 group 16-24 = 1.3%. Proportions not reported in B_II_: SOFA-2 score 4 = 1.7%. Proportions not reported in B_III_: SOFA-1 score 2 = 2.4%. Proportions not reported in B_IV_: SOFA-1 score 3 = 1.1%, SOFA-1 score 4 = 1.0%, SOFA-2 score 3 = 0.7% and SOFA-2 score 4 = 2.2%. Proportions not reported in B_V_: SOFA-1 score 3 = 0.4%, SOFA-1 score 4 = 0.0%, SOFA-2 score 3 = 0.3% and SOFA-2 score 4 = 0.0%. Proportions not reported in B_VI_: SOFA-1 score 3 = 1.6%, SOFA-1 score 4 = 0.1% and SOFA-2 score 4 = 1.8%. SOFA, sequential organ function assessment; MODS, multiorgan dysfunction syndrome.

### Association of organ dysfunction with ICU mortality

We next examined whether SOFA-2 organ scores had better stepwise associations with critical care outcomes than SOFA-1. Brain, respiratory, cardiovascular and renal domains showed some degree of stepwise progression of critical care mortality for both SOFA-1 and SOFA-2 systems (**Fig. 2A-D**). The respiratory, cardiovascular and renal components in SOFA-2 generated greater discrimination in critical care mortality rates at the higher ends of the scores, consistent with the aims of the SOFA reclassification. In contrast, the liver and haemostasis domains showed much flatter mortality profiles across the severities of organ dysfunction (**Fig. 2E-F**). These patterns were mirrored in vasopressor and ventilator days (**Supplemental Fig. 2 and 3**). Overall, SOFA-2 scoring appeared to describe stepwise increases in ICU mortality across the dominant domains of organ dysfunction seen in trauma patients.

**Figure 2.**
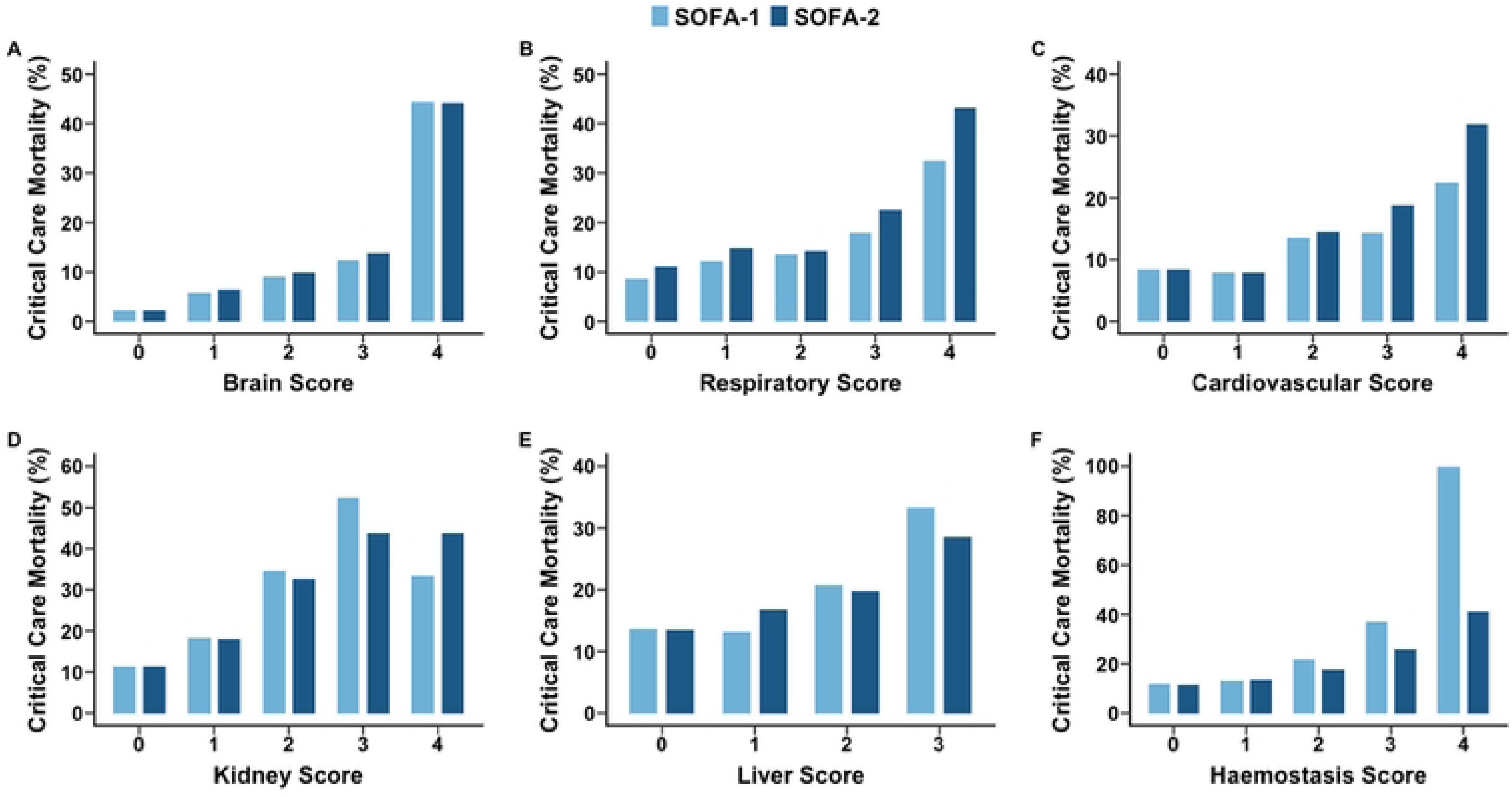
Critical care mortality according to individual SOFA-1 and SOFA-2 component scores (A-F). SOFA, sequential organ function assessment.

### Definition of MODS between SOFA1 and SOFA2

Given the differences between the scoring systems, we then examined changes in MODS classification across the two scores (**Table 2**). While most patients remained in their MODS/No MODS category in both scoring systems, 159 patients (7.3%) who were defined as having MODS by SOFA-1 were reclassified as No MODS by SOFA-2 (**Table 2**, **Fig. 3A**). Whilst injury severity was similar across all cohorts, this “downgraded” group (SOFA-1 MODS to SOFA-2 No MODS) had a significantly higher proportion of TBI than those classified as No MODS in both systems (42.1% vs 26.3%, <0.001). Critical care mortality was 7.5% in this group, whereas those classified as No MODS by both SOFA-1 and SOFA-2 had a mortality of 0.7% (p<0.001; **Table 2 and Fig. 3B**). The SOFA-1 MODS to SOFA-2 No MODS group also had substantially higher vasopressor and ventilator requirements (**Fig 3C-D**) as well as longer critical care and hospital stay than the No MODS group (**Table 2**). In summary, SOFA-2 reclassifies a group of SOFA-1 MODS patients as No MODS, and these trauma patients have meaningfully higher resource utilisation and mortality than the rest of the No MODS patients.

**Figure 3.**
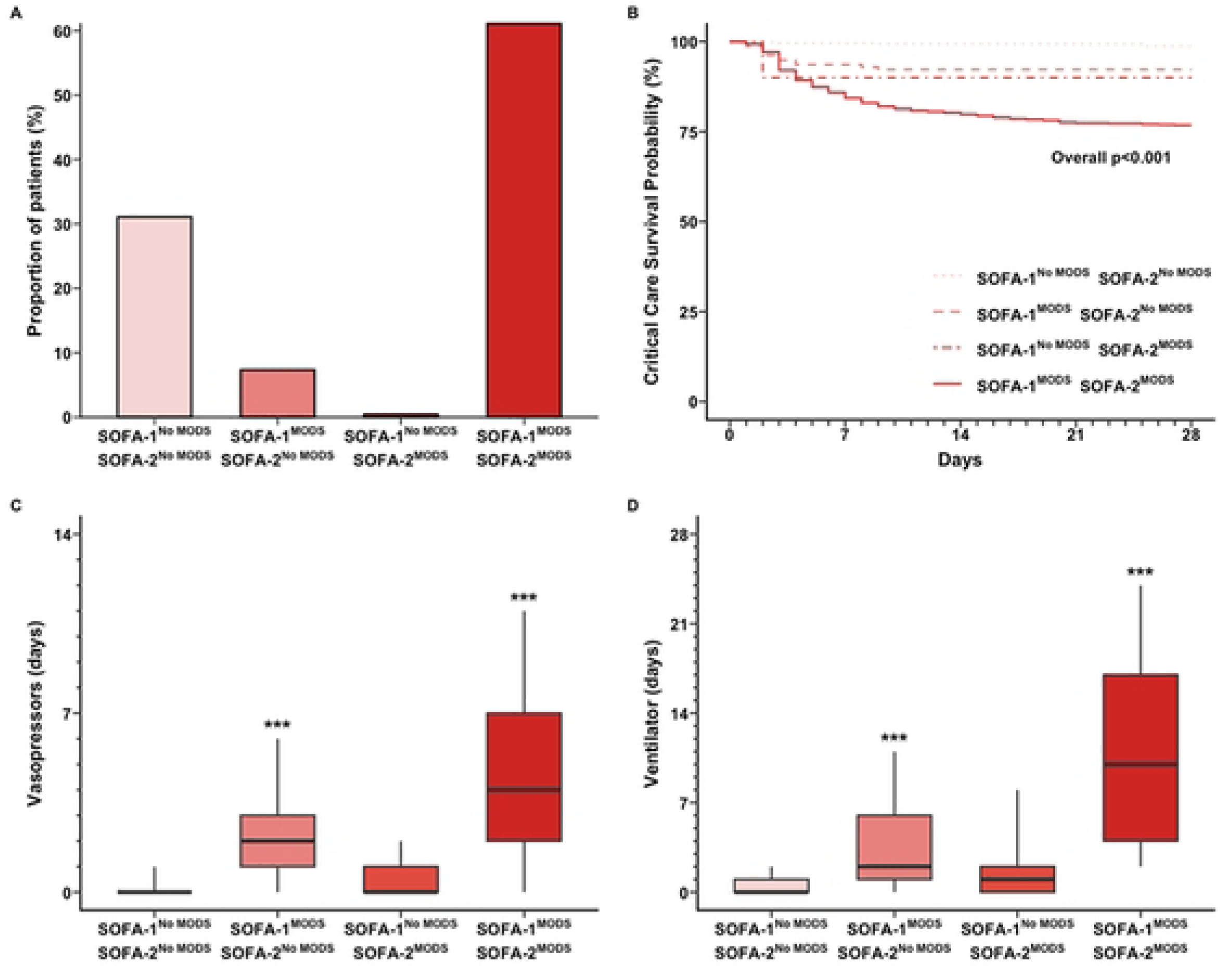
Reclassified groups and clinical outcomes (critical care survival, vasopressor days and ventilator days) according to SOFA-1 and SOFA-2. Kaplan-Meyer p value was derived using log-rank test. Kruskal-Wallis test with Dunn’s post-hoc was used to compare vasopressor days and ventilator days across groups. SOFA, sequential organ function assessment; MODS, multiorgan dysfunction syndrome.

**Table 2.** Characteristics of the study cohort according to MODS classification between SOFA-1 and SOFA-2.

|  | All patients<br>N = 2162 | SOFA-1<sup>No MODS</sup><br>SOFA-2<sup>No MODS</sup><br>N = 672 | SOFA-1<sup>MODS</sup><br>SOFA-2<sup>No MODS</sup><br>N = 159 | SOFA-1<sup>No MODS</sup><br>SOFA-2<sup>MODS</sup><br>N = 10 | SOFA-1<sup>MODS</sup><br>SOFA-2<sup>MODS</sup><br>N = 1321 |
| --- | --- | --- | --- | --- | --- |
| **Demographics** |  |  |  |  |  |
| Age, years | 46 (28-65) | 40 (25-61) | 46 (29-70) | 52 (45-62) | 48 (30-65)*** |
| Sex, male | 1659 (76.8%) | 506 (75.3%) | 116 (73.0%) | 7 (70.0%) | 1030 (78.0%) |
| **Clinical characteristics** |  |  |  |  |  |
| SBP, mmHg | 124 (100-145) | 125 (102-143) | 123 (106-148) | 133 (115-150) | 124 (97-147) |
| Glasgow coma score | 13 (6-15) | 15 (13-15) | 14 (9-15)** | 11 (8-14) | 9 (4-14)*** |
| Base deficit, mEq/L | 3.6 (1.0-7.1) | 2.6 (0.5-5.0) | 2.6 (-0.2-6.1) | 2.5 (-0.9-5.0) | 4.5 (1.7-8.3)*** |
| **Injury characteristics** |  |  |  |  |  |
| MOI, blunt | 1821 (84.2%) | 505 (75.1%) | 124 (78.0%) | 10 (100.0%) | 1182 (89.5%)*** |
| Traumatic brain injury | 997 (46.1%) | 177 (26.3%) | 67 (42.1%)*** | 4 (40.0%) | 749 (56.7%)*** |
| Injury severity score | 25 (17-34) | 25 (16-29) | 25 (16-34) | 23 (13-29) | 26 (18-38)*** |
| Head Neck AIS | 3 (0-5) | 1 (0-4) | 3 (0-4) | 3 (0-4) | 3 (0-5)*** |
| Face AIS | 0 (0-0) | 0 (0-0) | 0 (0-1)* | 1 (0-2) | 0 (0-1)*** |
| Thorax AIS | 3 (0-3) | 2 (0-3) | 2 (0-3) | 3 (0-4) | 3 (0-4) |
| Abdomen-Pelvis AIS | 0 (0-2) | 0 (0-2) | 0 (0-2) | 0 (0-2) | 0 (0-2)** |
| Extremity AIS | 0 (0-2) | 0 (0-2) | 0 (0-2) | 2 (0-3) | 0 (0-2) |
| External AIS | 0 (0-0) | 0 (0-0) | 0 (0-1) | 0 (0-0) | 0 (0-0) |
| **Transfusion requirements** |  |  |  |  |  |
| MHP Activation | 765 (35.7%) | 178 (26.6%) | 46 (29.3%) | 1 (10.0%) | 540 (41.3%)*** |
| Red blood cells in 24hr | 0 (0-4) | 0 (0-2) | 0 (0-4) | 0 (0-1) | 1 (0-5)*** |
| **Clinical Outcomes** |  |  |  |  |  |
| Critical care mortality | 314 (14.5%) | 5 (0.7%) | 12 (7.5%)*** | 1 (10.0%)* | 296 (22.4%)*** |
| Critical care length of stay | 9 (4-18) | 4 (2-6) | 7 (4-13)*** | 12 (4-16)* | 16 (9-24)*** |
| Mortality | 369 (17.1%) | 16 (2.4%) | 13 (8.2%)** | 1 (10.0%) | 339 (25.7%)*** |
| Length of stay | 27 (14-45) | 14 (8-24) | 19 (13-33)*** | 19 (12-38) | 37 (25-55)*** |

Data is presented as median (interquartile range) or frequency (percentage). Continuous variables were compared using Kruskal-Wallis rank sum test with Dunn's post-hoc test. Categorical variables were compared using Fisher's exact test. Length of stay reported for survivors only.
Compared to SOFA-1<sup>No MODS</sup> SOFA-2<sup>No MODS</sup>: \*, p<0.05; \*\*, p<0.01; \*\*\*, p<0.001.
MODS, multiorgan dysfunction syndrome; SOFA, sequential organ function assessment; SBP, systolic blood pressure; MOI, mechanism of injury; AIS, abbreviated injury score; MHP, major haemorrhage protocol.

## Discussion and conclusions

In this multicentre study of over 2000 severely injured patients, the updated SOFA-2 scoring system resulted in a lower prevalence of multiorgan dysfunction syndrome compared with the original SOFA-1 score. The reclassification in scores was predominantly driven by changes in the respiratory and cardiovascular domains. Increasing SOFA-2 scores were associated with stepwise increases in ICU mortality, especially in the dominant domains of trauma-related organ dysfunction. Finally, we found that SOFA-2 downgraded a group of severely injured trauma patients with high mortality rates to No MODS status. This clinically meaningful variation warrants consideration in future studies of organ dysfunction in trauma.

Our study is the first to specifically evaluate the SOFA-2 score in a severely injured trauma population. MODS following severe traumatic injury develops as a result of complex inter-relationships between widespread inflammation and immune dysregulation, together with metabolic derangements (22,23). As contemporary early trauma resuscitation has evolved, more trauma patients survive the initial phases of management, yet MODS in critical care continues to be associated with high mortality rates and resource use (24). We found that respiratory, cardiovascular and renal dysfunction in SOFA-2 were better discriminators of critical care mortality, consistent with the results of a recent population-level study illustrating good mortality discrimination across a wide range of patient groups despite significant heterogeneity in pathophysiology(1(25)). Our work illustrates that SOFA-2 reclassification is an important practical consideration in future studies of organ injury in trauma patients, most notably in relation to the prevalence of MODS and respiratory dysfunction/failure. This has implications for clinical and mechanistic phenotyping studies in trauma, as well as selection criteria and outcome measures in future clinical trials that focus on targeted disease-modifying interventions for organ protection.

We found that application of SOFA-2 reduced the percentage of trauma patients meeting criteria for MODS and differences between scoring systems were driven largely by changes in individual organ components. The most pronounced divergence was observed in respiratory scoring, where SOFA-2 classified a substantially larger proportion of patients as having no respiratory dysfunction at 24 hours. This is likely to be attributable to changes in cut offs in the new SOFA-2 system, which lower the P/F ratio required to score 0 from 53 to 40 kPa (400 to 300 mmHg). Although intubation and ventilation is common in trauma cohorts, our results indicate that only 10% had significant respiratory compromise according to SOFA-2, and thus trials using endpoints focusing solely on respiratory failure in trauma risk being underpowered. Cardiovascular scores were more evenly distributed early after injury under SOFA-2 compared to SOFA-1. This is likely due to updated scoring criteria reflecting increased use of noradrenaline as a first line vasopressor therapy (26), alongside more balanced scoring thresholds. The highest levels of renal dysfunction were more frequently identified under SOFA-2, which likely reflects the use of renal replacement therapy (RRT) as the criteria required to score 4. This should be cautiously interpreted, as other factors may impact RRT utilisation beyond levels of renal dysfunction. For example, timing of initiation of RRT differs between clinicians and remains controversial (27,28), and may not correspond to acute kidney injury (AKI) stages previously utilised for scoring.

For the majority of our cohort the MODS or No MODS classification did not change, although there were important differences in those who were reclassified. We identified a small but clinically significant group who in SOFA-2 were classified as No MODS but have high mortality and organ support requirements. The precise reasons that this ‘occult MODS’ group of injured patients did not reach the SOFA-2 threshold for MODS is not clear. This group had a high incidence of TBI but no other obvious differences in clinical characteristics. Whilst brain injury is thought to potentially confound the assessment of MODS (9,29), complex interactions between significant systemic inflammation and CNS injury have been linked to MODS development in TBI patients (30,31). The presence of MODS is known to worsen outcomes for those with severe TBI (31,32) but within this reclassified group it is currently unclear whether this reflects brain injury driven inflammation or organ support needs for patients with severe TBI. Contemporary organ dysfunction in multiply injured patients with concomitant severe TBI requires further investigation.

This study has some limitations. We combined the data from two studies and there were minor differences in collection. Namely, in the MODET study, renal replacement therapy was only recorded for 21 of the 28 days, so this may be an underrepresentation of renal dysfunction. Secondly, we did not conduct any imputation for missing data. We used carry-forward methodology previously described in trauma studies and in the development of SOFA-2 (5,33), which assumes that data not recorded is the same as the previous day. This may affect organ dysfunction trajectories, but we excluded cases with missing SOFA data for the MODS analysis. Thirdly, for most patients in ACIT-II and MODET, GCS was recorded as a complete score, therefore we could not use the GCS motor component score for CNS scoring as described in SOFA-2 methodology. Finally, trauma critical care mortality is associated with many factors other than MODS and we did not adjust for this, however mortality causation was not an objective of this study.

In conclusion, SOFA-2 reduces trauma MODS prevalence and generates substantial changes in organ dysfunction classification, particularly in respiratory and cardiovascular domains. This represents an important update in the measurement of trauma MODS and better reflects contemporary trauma critical care. However, SOFA-2 reclassified a small cohort from MODS to No MODS, with outcomes more consistent with MODS. Future studies should validate SOFA-2 thresholds in large-scale trauma populations, with a focus on investigating multiply injured patient cohorts with traumatic brain injuries.

## Data Availability

An anonymised dataset has been uploaded as a supporting information file to this submission. Further information can be obtained by contacting the corresponding author.

## Acknowledgements

We acknowledge the work of the ACIT-II and MODET study group and authors for the previous work and recruitment of patients upon which this study was based.

## Supplemental Figure Captions

Supplementary Figure 1. Flow diagram of the study cohort. ACIT-II, Activation of Coagulation and Inflammation in Trauma II observational study; MODET, Multiple Organ Dysfunction in Elderly Trauma study; SOFA, sequential organ function assessment; ISS, injury severity score.

Supplementary Figure 2. Vasopressor days in survivors according to individual SOFA-1 and SOFA-2 component scores. No survivors in the SOFA-2 Haemostasis Score = 4. SOFA, sequential organ function assessment.

Supplementary Figure 3. Ventilator days in survivors according to individual SOFA-1 and SOFA-2 component scores. No survivors in the SOFA-2 Haemostasis Score = 4. SOFA, sequential organ function assessment.

## Notes

### Competing Interest Statement

The authors have declared no competing interest.

### Author Declarations

Patients were included if they had been enrolled in the Activation of Coagulation and Inflammation in Trauma II observational study (ACIT-II, NHS Research Ethics 104 Committee: 07/Q0603/29) (15) between 2011 and 2024 or in the Multiple Organ Dysfunction in Elderly Trauma study (MODET, NHS Research Ethics Committee: 106 16/LO/1720)

